# Genetic associations of risk behaviours and educational attainment

**DOI:** 10.1101/2023.04.24.23289036

**Authors:** Michelle Arellano Spano, Tim Morris, Neil M Davies, Amanda Hughes

## Abstract

Risk behaviours such as alcohol use, smoking, and physical inactivity are common in adolescence and persist into adulthood. People who engage in more risk behaviour are more likely to have lower educational attainment. Genome-wide association studies show that participation in risk behaviours and level of education are both heritable and have a highly polygenic architecture, suggesting an important role of many variants across the genome. The extent to which risk behaviours and educational attainment have shared genetic overlap is unknown, yet knowledge of this could help understand how these traits co-occur and influence each other. In the ALSPAC cohort, we used genome-based restricted maximum likelihood (GREML) to estimate the genetic covariance between risk behaviours and educational achievement. We found a strong genetic component of educational achievement and a modest genetic component of the risk behaviours. Whereby children who have a higher genetic liability for education also have a lower genetic liability for risky behaviours.

## Introduction

Adolescence is a crucial formative period for an individual’s future well-being; the choices made during this period can have important repercussions later in life. (1) Risk behaviours like alcohol use, smoking and physical inactivity are often first engaged in during adolescence and persist into adulthood. (2) These behaviours can influence the development of noncommunicable diseases (NCDs) later in life and increase the risk of injury, substance dependence, morbidity, and low educational attainment. For example, studies have shown that those who smoke, drink excessively and have an unhealthy diet and lifestyle are at a higher risk of developing cardiovascular disease, type 2 diabetes, and cancer. (3,4) Likewise, engaging in multiple risk behaviours is associated with a greater risk of developing chronic diseases and increased mortality. (5) Individuals who engage in risk behaviours are more likely to present lower educational achievement than those who engage in fewer risk behaviours. (6)

Risk behaviours tend to cluster within individuals. Schuit et al. (7) found that at least 25% of adults report currently engaging in three or more risk behaviours. This clustering can occur because of various reasons. First, engagement in one behaviour can lead to engagement in other risk behaviours, in a process known as co-occurrence. (8) For example, alcohol use can increase the risk of engaging in other risk-taking behaviours, such as risky sexual behaviour, via inhibition mechanisms affecting an individual’s decision-making processes. (9) The aforementioned effect, where one behaviour causes the other was also exampled by Bellis et al. (10) that found that early substance use is associated with an increased risk of engaging in premature sexual activity in adolescent girls. Second, features of an adolescent’s social and psychological environment, such as peers’ behaviour, can influence engagement in risk behaviours. For example, an individual is more likely to engage in risk behaviours if people around them engage in risk behaviours. (11) If environmental influences affect multiple risk behaviours at the same time, this can result in clustering of risk behaviours together in a process of environmental confounding. One source of environmental confounding are dynastic effects, which occurs when parent’s heritable traits affect the child’s trait. This is particularly evident in genetic studies of intergenerational transmission of education; where genetic traits associated with educational achievement in a parent’s generation may lead to the creation of educationally rich environment. (12) In turn, this will have an impact on the children educational achievement via genotypic and phenotypic inheritance. Ultimately inducing correlation through confounding between genotypes and phenotypes. These processes, in terms of the scope of this research are illustrated in figure 1, where the focus is the relationship of clustered risk behaviours – clustered by processes of co-occurrence and environmental confounder - and educational achievement.

**Figure 1:**
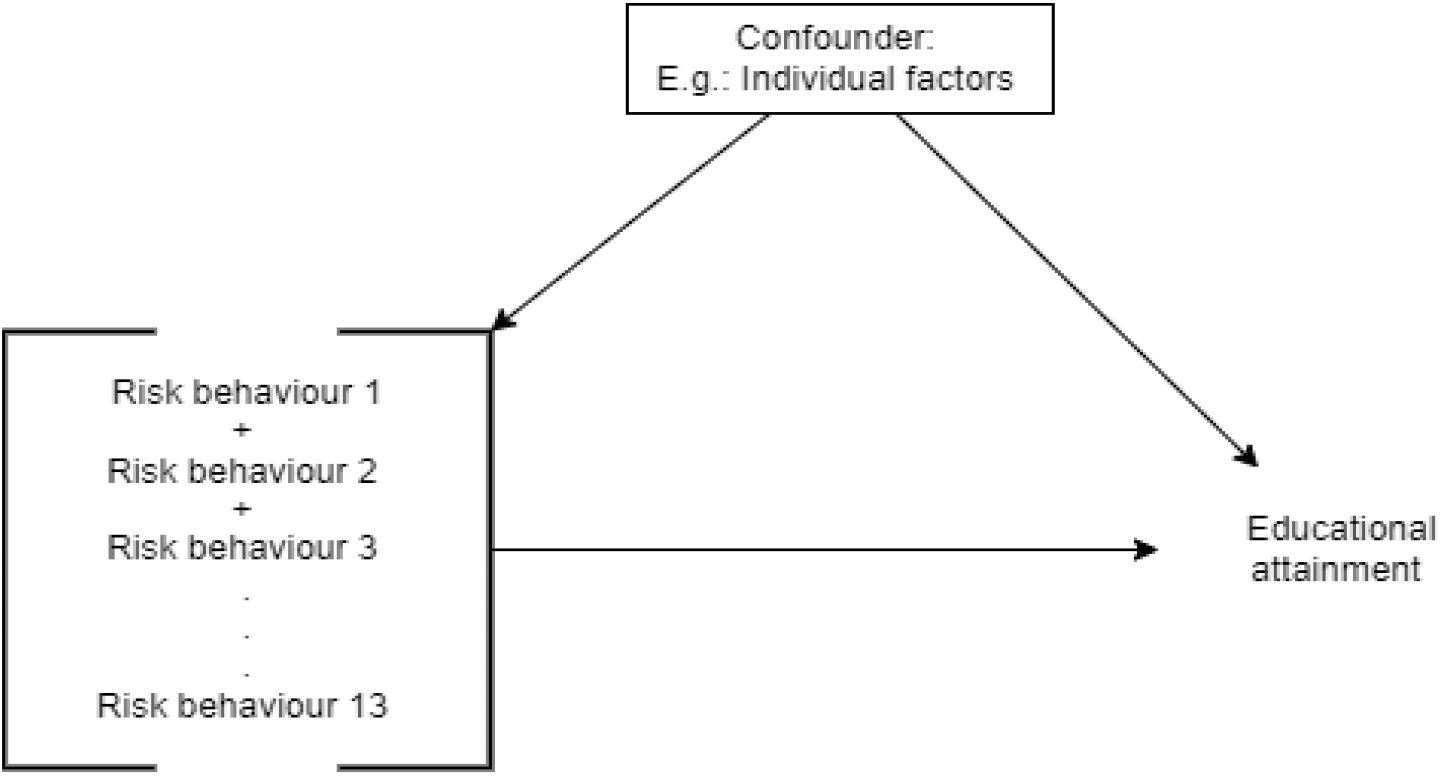
Directed acyclic graph for the relation between multiple risk behaviours and educational achievement.

Observational studies have focused on the effect of risk behaviours on various behavioural and social outcomes throughout time. These report associations between risk behaviours in adolescence and socioeconomic position later in life (13), adult aggression (14) and continuity of substance misuse. (15) Likewise, a medical survey found in a sample of 3,059 Canadian students that adolescent engagement in risk behaviours is associated with concussions, indicating engagement in risk behaviours as an etiological component of adolescent concussions. (16) Wright et al. (6) found an association between multiple risk behaviours and educational achievement, where a adolescent’s engagement in each additional risk behaviour was associated with 23% lower odds of attaining five A*-C grades at age 16. Risk behaviours in adolescence, if causal, could therefore be a key target for interventions aiming to improve socioeconomic as well as health outcomes. This is supported by a systematic review of interventions targeting multiple risk behaviours, which found that school-based intervention may play an important role in preventing adolescence engagement in tobacco, alcohol and illicit drug use and improving physical activity in adolescence. (17) However, it is unclear whether risk behaviours causally affect educational achievement, as opposed to both being influenced by features of the environment (confounding) or educational achievement influencing risk behaviours (reverse causation). Genetically informed studies can help overcome these sources of bias and improve our understanding of the causal relationships between education and risk behaviours in adolescents. In the presented research, we used genetics methods to investigate the genetic architecture of risk behaviours and educational achievement, and the effects that risk behaviours have on educational outcomes and vice versa.(18)

## Methods

### Study participants

The Avon Longitudinal Study of Parents and Children (ALSPAC) is a prospective birth cohort based in the Bristol and Avon area in the UK. Pregnant women were invited to take part if they were resident in the area and had expected dates of delivery 1st April 1991 to 31st December 1992. From 14,541 pregnancies initially enrolled, 13,988 children were alive at 1 year of age. When the oldest children were approximately 7 years of age, an attempt was made to include eligible cases who had not joined the study originally. The total sample size for analyses using any data collected after the age of seven is therefore 15,447 pregnancies, resulting in 15,658 foetuses. Of these 14,901 children were alive at 1 year of age. Data collected from age seven onwards is therefore available for 14,901 participants, with details of the enrolment phases provided elsewhere. (19–21) We carried out our GREML analyses in a complete case sample composed of participants with genetic information and all information on covariates and risk behaviours. The complete case sample was derived from the ALSPAC cohort, starting with 15,616 participants. This sample was then restricted to those who had genetic data available and were unrelated (N=8,815). Of the genetic sample, 2,366 participants had information on some risk behaviours. Finally, our analytical sample was composed of 1,735 who had complete genetic data, risk behaviours and full information on covariates (see supplementary Figure 1 for STROBE diagram). We carried out the phenotypic analyses in an imputed sample. In order to maintain the sample size, we imputed the missing values by chained equations in Stata. During the multiple imputation 50 datasets were created. Besides variables used in the analysis, we included marital status, mother’s smoking status, maternal education, housing tenure and parental social class as auxiliary variables. We used logistic regression to impute the risk behaviours, linear and truncated regression for continuous variables and order logistic regression to impute categorical variables.

Ethical approval for the study was obtained from the ALSPAC Ethics and Law Committee and the Local Research Ethics Committees. Consent for biological samples has been collected in accordance with the Human Tissue Act (2004) (for full information on ALSPAC ethical approval please see: http://www.bristol.ac.uk/alspac/researchers/research-ethics/). Informed consent for the use of data collected via questionnaires and clinics was obtained from participants following the recommendations of the ALSPAC Ethics and Law Committee at the time. Ethical approval for the study was obtained from the ALSPAC Law and Ethics committee and local research ethics committees (NHS Haydock REC: 10/H1010/70). Completion of individual questionnaires was taken as consent for use of data from that questionnaire, with additional written consent from parents for use of clinic data. At age 16, young people and their parents gave written informed consent for use of the young person’s genetic information. At age 18, study children were sent ‘fair processing’ materials describing ALSPAC’s intended use of their health and administrative records and were given clear means to consent or object via a written form. Education data were not extracted for participants who objected, or who were not sent fair processing materials. (19,22)

### Genotyping

The genotypic data used in this analysis comes from the ALSPAC cohort and was obtained from blood, cell line and mouthwash samples, then genotyped using reference panel and standard quality control approaches. (14) ALSPAC children were genotyped using the Illumina HumanHap550 platform, and standard quality control procedures applied. Individuals were excluded for gender mismatches, minimal or excessive heterozygosity, disproportionate individual missingness (>3%) and insufficient sample replication (IBD < 0.8). During genetic quality controls individuals with non-European ancestry were removed, which is standard practice in genetic studies to minimize bias due to ancestral population stratification. SNPs with a minor allele frequency of <1%, call rate of <95% or evidence of Hardy–Weinberg disequilibrium 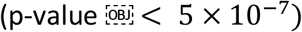. Cryptic relatedness was measured as proportion of identity by descent (IBD > 0.1). Imputation was performed using Impute v2.2.2 to the Haplotype Reference Consortium (HRC) panel and SNPs with poor imputation quality (info score < 0.8) removed.

### Measures

#### Multiple Risk Behaviours (MRBs) at age 16

An index of multiple risk behaviours (MRBs) was derived from two main data collections during the participants’ adolescence: a self-completed questionnaire issued during a clinic assessment at age 15 and a self-completed postal questionnaire at age 16. We coded 13 risk behaviours into binary format (no=0; yes=1) following Wright et al. (6) and then calculated an MRB index as the total number of risk behaviours each participant had engaged in. The study website contains details of available data through a searchable data dictionary and variable search tool: http://www.bristol.ac.uk/alspac/researchers/our-data/.

The risk behaviours included in the index were:

Physical inactivity: Participant has typically exercised <5 times per week over the past year.

TV Viewing: Participant spent three or more hours watching TV on average per day across the week.

Car passenger risk: Participant had been in a car passenger at least once in their lifetime where the driver (1) had consumed alcohol or (2) did not have a valid licence, or (3) the participant chose not to wear a seat belt last time travelling in a car, van, or taxi.

Scooter risk: Participant reported that they had last ridden a scooter within the previous four weeks and had not used a helmet on the most recent occasions.

Cycle helmet use: If the participant reported that they had last ridden a bicycle within the previous four weeks and they had not used a helmet on the most recent occasion.

Illicit drug use/solvent use: In the year since their 15th birthday, participant had either been a regular user (used more than five times) of one or more illicit drugs (excluding cannabis), including amphetamines, ecstasy, lysergic acid diethylamide (LSD), cocaine, ketamine or inhalants including aerosols, gas, solvents, and poppers.

Cannabis use: Participant who reported using cannabis ‘sometime, but less often than once a week’ or more regular use were classified as occasional users.

Regular tobacco use: Participant has ever smoked and is regularly smoking at least one cigarette per week.

Hazardous alcohol consumption: In the past year, participant had scored eight or more on the Alcohol Use Disorders Identification Test (AUDIT), indicating hazardous alcohol consumption.

Self-harm: Participant said they had purposely hurt themselves in some way in their lifetime.

Penetrative sex before the age of 16: Participant reported having had penetrative sex in the preceding year and that they were under 16 at the time.

Unprotected sex: Participant engaged in penetrative sex without using contraception on the last occasion they had had sex in the past year.

Criminal behaviour: Participant reported that at least once in the past year, they had undertaken at least one of the following seven offences: carried a weapon; physically hurt someone on purpose; stolen something; sold illicit substances to another person; damaged property belonging to someone else either by using graffiti, setting fire to it, or destroying or damaging it in another fashion; subjected someone to verbal or physical racial abuse; or been rude/rowdy in a public place.

As each of the risk behaviours can be represented as a binary indicator (see table 1 for descriptives of individual risk behaviours), we can denote the variable measuring engagement in risk behaviour *j* for each individual *i* by the binary indicator as follows:

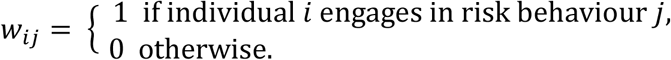

**Table 1:**
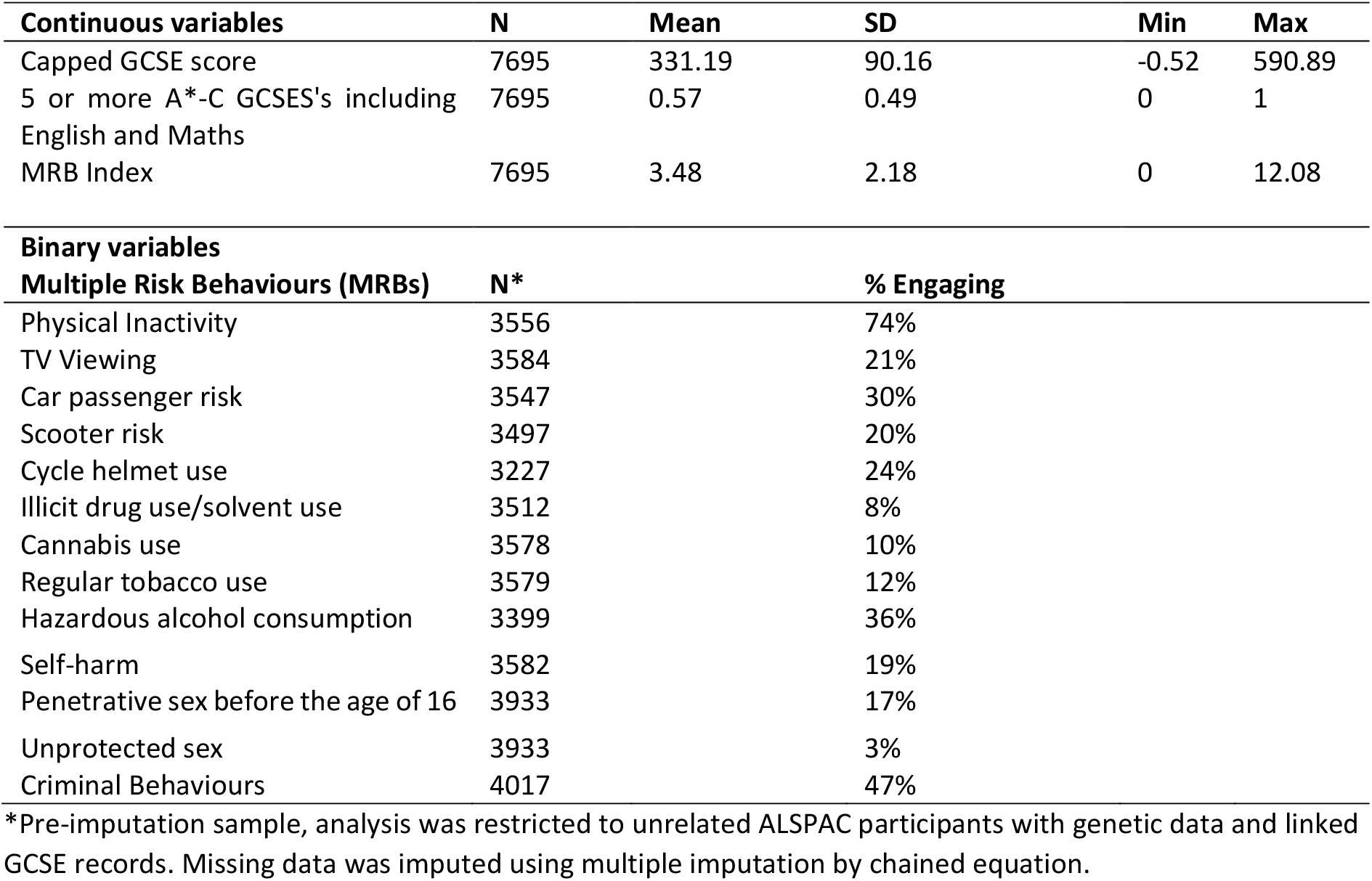
Descriptive statistics for education outcomes and MRB index in the imputed sample and individual multiple risk behaviours in the pre-imputed set.

Since we are looking at the overall engagement across a range of risk behaviours rather the individual effects of each, we then create a new single variable called the Multiple Risk Behaviour Index (MRBI), defined for each individual *i* as the sum of all behaviours, as follows:

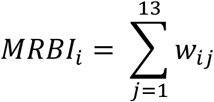

The new regressor *MRBI*_*i*_ is our exposure of interest summarised in Table 1.

### Educational achievement

Information on educational achievement was obtained via record linkage to the National Pupil Database (NPD). Managed by the Department of Education in England, it includes data collected from students in state-funded education and higher education from 2 to 21 years. This comprises the most complete and accurate record of compulsory education available in England. Educational measures were based on participants’ General Certificate of Secondary Education (GCSE) qualifications which are taken during educational Key Stage 4 when pupils are aged between 14 and 16 years old. At the time, Key Stage 4 marked the end of compulsory education in England. For this analysis we used two measures of achievement. The first was the capped GCSE score, which sums the student’s 8 best grades to obtain a summary measure of general achievement commonly used in educational research. Individual GCSE qualifications in each subject contribute 58 points for an A* through to 16 points for a G and 0 for a U (ungraded). Our second measure of educational achievement was whether participants achieved five or more A*-C grades at GCSEs. We used this as it is a minimum qualification requirement for entry to post-16 education and training courses.

### Polygenic Index (PGI)

Polygenic indexes for risk and education were calculated using genome-wide-significant single-nucleotide polymorphisms (SNPs) previously associated in GWAS with each trait at p<5.× 0*x*10^−8^. First, the risk PGI were calculated using the Karlsson Linner et al. (23) GWAS on risk behaviour and weighted using the individual SNP-coefficient from the GWAS for both mother and children. The full GWAS results includes the top hits extracted from UKBiobank and 23andMe, we excluded those SNPS that were not available in the ALSPAC sample and used the MRinstruments R package to identify the SNPs independently associated with risk. The clumping threshold was of 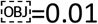 and 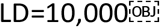 at 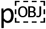. Then, we calculated a risk polygenic index for mothers and children using Plink 1.9. *r*^2^ Likewise, we calculated the education PGI for mothers and children using the largest GWAS for education, 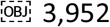 approximately uncorrelated 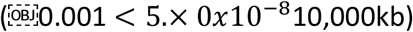 SNPS highly associated with educational attainment 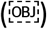. The predictive power of the educational achievement PGI, as measured by 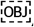, was 10% for the Capped GCSE Score (continuous outcome) and *r*^2^ = for the children’s PGI. For mothers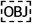, the reported 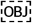 was of 7% and 4% for the continuous and binary outcome *R*^2^ For the risk behaviours, the 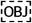 for children’s risk PGI were of *R*^2^ and for mothers’ risk PGI 0.1%.

### Statistical analysis

In order to explore the association between the MRB index and educational achievement, we carried out three types of analyses. First, we did a phenotypic analysis on the imputed sample to interrogate the association between MRB index and the continuous and binary measures for educational attainment in the ALSPAC cohort. Secondly, in order to explore the genetic aetiology of engagement in risk behaviour and educational achievement we performed a GREML analysis to explore the heritability of this traits, this analysis was carried out in the complete case sample. Lastly, given the possible bias that are present in observation causal association we explore the directionality of the effect employing a MR and bidirectional MR analyses in our imputed datasets. The aforementioned analyses and respective results are further explained below.

#### A) Phenotypic associations

We used linear and logistic regression to estimate the association of the MRB Index and capped GCSE score and gaining five or more GCSE grade A*-C respectively. Our baseline results were only adjusted for the young person’s sex. Since it is possible that other factors may confound the association of educational achievement and the number of risk behaviours, we also estimated these associations adjusted for potential confounders linked to socioeconomic position and cognitive ability. These estimates were adjusted for parental social class, maternal education, and housing tenure at the time of the child’s birth. Lastly, we estimated a third set of associations adjusted for sex, socioeconomic factors, and the child’s cognitive ability. Table 2 shows the different categories used for each of these controls respectively.

**Table 2:** Summary statistic of parental social class, housing tenure, maternal education, and participants’ sex.

| ***Variables*** | ***N*** | ***Mean*** | ***Cum.*** |
| --- | --- | --- | --- |
| **Parental Social Class (N=7,695)** |  |  |  |
| Professional | 1,113 | 14.5 | 14.5 |
| Managerial and technical | 3,316 | 43.4 | 57.9 |
| Skilled non-manual | 1,929 | 25.2 | 83.1 |
| Skilled manual | 923 | 12 | 94.63 |
| Partially unskilled | 365 | 4.74 | 99.37 |
| Unskilled | 48 | 0.63 | 100 |
| **Housing Tenure (N=7,695)** |  |  |  |
| Owned | 6,127 | 79.6 | 78.6 |
| Council rented | 782 | 10.2 | 89.8 |
| Privately rented | 786 | 10.2 | 100 |
| **Maternal Education (N=7,695)** |  |  |  |
| <O level | 1,943 | 25.3 | 25.3 |
| O level | 2,674 | 34.8 | 60 |
| A level | 1,897 | 24.7 | 84.6 |
| Degree | 1,182 | 15.4 | 100 |
| **Participant's Sex (N=7,695)** |  |  |  |
| Female | 3,945 | 51.3 | 51.3 |
| Male | 3,750 | 48.7 | 100 |

#### B) Bidirectional Mendelian Randomization (MR)

Mendelian Randomization (MR) is a statistical method that uses genetic variants as instrumental variables to test causal effects between a purported exposure and an outcome in observational data. Mendelian randomization uses the fact that alleles are randomly transmitted from parents to offspring during conception (24) This method can evaluate the causal relationships by using genetics variants as instrumental variables for the exposure to estimate causal effects from exposures to outcomes. Since the genetics variants associated with the exposure do not change in response to a person’s health or environmental circumstances, associations between exposure-associated genetic variants and the outcome are not affected by classical cofounding or reverse causation that often affect observation studies estimates. (25) For MR estimates to be valid, the genetic instruments must meet three assumptions: 1) relevance, it must associate with the exposure, 2) independence, there must be nothing that causes both the instrument and the outcome, and 3) exclusion, the association of the instrument and the outcome must be entirely mediated via the exposure. (26)

For educational and risk behaviours, a causal effect in either direction is plausible, so we used bidirectional Mendelian Randomization. Bidirectional Mendelian Randomization is an extension of a standard MR analysis which attempts to differentiate whether the exposure is a cause of the outcome, a consequence of the outcome, or if there is a true bidirectional causal effect between them. (27)

First, we used MR to estimate the effect of educational achievement on risk behaviours. We used a two-stage least squares instrumental variable model (Stata’s ivreg2) with the risk behaviours index as the outcome and instrumented educational attainment using a polygenic score of SNPs previously associated with years of schooling. (28) Next, we used MR to estimate the effect of risk behaviours on educational achievement. We used a two-stage least squares instrumental variable model (Stata’s ivreg2) with capped GCSE points score as the outcome and instrumented the risk behaviours index using a polygenic score of SNPs previously associated with risk-taking behaviour. (23) For each outcome, two sets of models were run: one which adjusted for the young person’s sex and their first 20 principal components of ancestry, and one which additionally adjusted for the mother’s polygenic score. Likewise, for the binary outcome obtaining 5 or more A*-C GCSEs, we used a two-stage least squares instrumental variable model, and again instrumented the risk behaviours index using a polygenic score of SNPs previously associated with risk-taking behaviour.

#### B) SNP Heritability and bivariate heritability

We conducted genomic-based restricted maximum likelihood (GREML) to examine the genetic overlap between MRB Index and educational achievement. These models were carried out using Genome-wide Trait Analysis (GCTA). GCTA uses a genomic restricted maximum likelihood (GREML) method to estimate the proportion of phenotypic variance that is statistically explained by all measured genome-wide single nucleotide polymorphisms (SNPs) for complex traits, known as the SNP-based heritability. GCTA estimates heritability by comparing the genetic similarity of unrelated individuals to their phenotypic similarities. Unrelated participants (defined as more distantly related than second cousins) are determined using the ALSPAC Genetic Relatedness Matrices (GRMs). (29) If a phenotype is explained by the genetic variants, then we would expect more genetically similar individuals to be more phenotypically similar. We first carried a univariate model to test the educational outcomes and MRB Index, specified as:

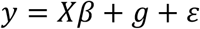

where *y* is the heritability of the phenotype (MRB Index), *X* is a series of covariates, *g* is a normally distributed random effect with variance 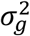 and *ε* is a residual error with variance 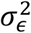. The SNP-based heritability, as the phenotypic variance measured by common genetic variance is:

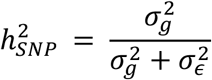

To control for population-specific variation in allele distribution that could potentially bias the estimate, the first 20 principal components of inferred population structure are included in the analysis as covariates.

We estimated genetic correlations using bivariate GCTA, by running set of analyses between the MRB Index and both measures of educational achievement, capped GCSE score and obtaining 5 or more A*-C GCSEs. Genetic correlations allow us to quantify the overlap in SNPs that are associated with one phenotype are also associated with others. Specifically for this study, the genetic correlation shows us the SNPs that are associated with MRB Index and are also associated with our measures of educational achievement. Thus, correlations of all genetic effects across the genome for a given phenotype A and B is estimated as

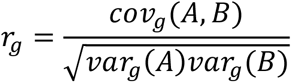

where *r*_*g*_ is the genetic correlation between phenotypes *A* and *B, var*_*g*_(*A*) is the genetic variance of phenotype *A*, and *cov*_*g*_(*A, B*) is the genetic covariance between phenotypes *A* and *B*. Genetic correlations can show common genetic architecture, where two phenotypes are influenced by the same SNPs. GCTA does not support GREML using multiply imputed phenotype data, so these analyses were performed in the subset of the sample used for the phenotypic and Mendelian randomization models who had complete phenotypic information (N=1735).

## Results

### Sample descriptive

Our starting sample was composed of 15,645 pregnancies, which was then restricted to those with genetic data available (N=8,815). Of this initial sample 51% were male and 49% female, and 7,496 had been successfully linked to their educational outcomes. Of these, 2,366 had information on some and all risk behaviours respectively and also had some information on covariates. Thus, 1,735 participants had no missing value. Table 2 in the supplementary shows the difference between our analytical sample, that had been restricted to those participants that had genetic information, all covariates and information on risk behaviour (N=1735) and the core ALSPAC sample (N=15,616). Based on the analytical sample, imputed the sample, and carried out the phenotypic analysis and MR analyses on the imputed sample (N=7,695).

### Phenotypic associations of risk behaviours and educational achievement

Tables 3 reports results from models which regress the capped GCSE score on the MRB Index using imputed data. The first column shows the results of the regression of the capped GCSE score on the MRB Index adjusted only for sex. A standard deviation increase in the MRB Index was phenotypically associated with a -0.14 (95% CI: [-0.17, -0.12]) standard deviation decrease in capped GCSE score. After adjusting for sex, parental socioeconomic position and maternal education, a standard deviation increase in the MRB Index corresponds to a -0.12 (95% CI: [-0.14, -0.10]) standard deviation decrease in capped GCSE score. This suggests that each additional risk behaviour is associated with lower capped GCSE scores. Likewise, results for the fully adjusted binary outcome model suggested the odds of obtaining five or more A*-C GCSEs were 19% (95% CI: [16%, 23%]) lower per extra risk behaviour (see supplementary, table 6).

**Table 3:**
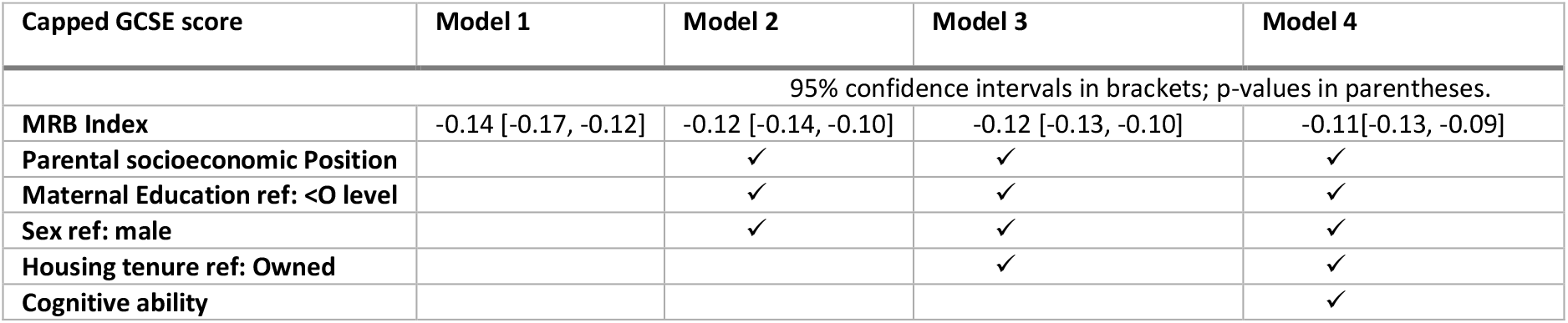
Associations of capped GCSE score with an index of multiple risk behaviours, based on imputed data (N=7,695)

**Table 4:** GCTA estimates. *Logistic regression, *h*^2^: univariate heritability, *r*_*g*_: genetic correlation.

| SNP-based (GCTA) estimates |  |  |  |  |
| --- | --- | --- | --- | --- |
| *Univariate estimates* | n | $h^2$ | SE | p |
| *Capped GCSE score* | 6646 | 0.55 | 0.05 | <7.9E12 |
| *MRBI* | 2080 | 0.21 | 0.16 | 0.089 |
| *Bivariate estimates* | n | $r_g$ | SE | p |
| *Capped GCSE score: MRBI* | 4363 | -0.49 | 0.27 | 0.005 |

### GREML

The univariate models show the association between the phenotypes of interest and the genotypic data. We observed SNP heritability in the educational outcome of 0.55 (p=<0.001) for capped GCSE scores. Likewise, we observed a moderate heritability estimate of the MRB Index of 0.21 (p=0.089) These results suggest that the variation in educational achievement and some of the risk behaviour score can be explained by common genetic variation.

The bivariate analysis shows a strong negative genetic correlation between the MRB index and educational outcomes of -0.49 (p=0.005) for capped GCSE score. This suggests considerable genetic overlap between these traits and that genetic variation associated with risk behaviours is also associated with lower educational achievement.

### Bidirectional Mendelian randomization

Figure 4 shows associations between the young person’s genetically-instrumented risk behaviour index and capped GCSE points score. There was little evidence of an impact genetically-instrumented risk behaviours on capped GCSE score when adjusted for the young person’s sex and principal ancestry components (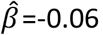, 95% CI: [-0.27,0.15]) or when additionally adjusted for the maternal risk PGI (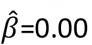, 95% CI: [-0.24,0.24]). The results for the binary outcome were similar (Supplementary figure 2). the case when the outcome was for GCSE capped points score, there was little evidence from that risk behaviours influenced educational achievement adjusted for maternal risk PGI (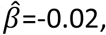, 95% CI: [-0.14,0.10]).

**Figure 3:**
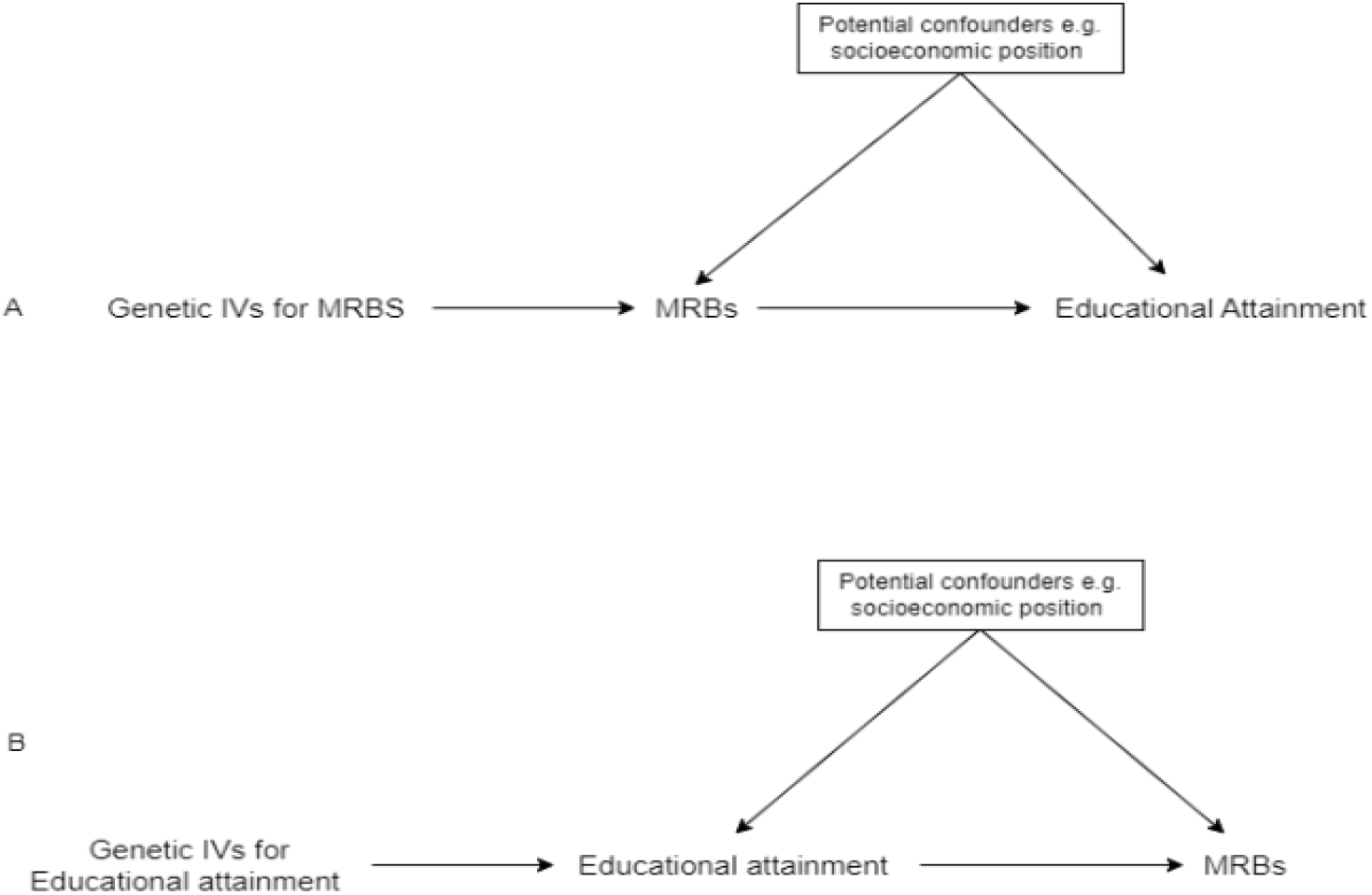
Directed acyclic graph of a bidirectional MR presenting the relationship between MRB Index and educational achievement. PGI: Polygenic scores. MRB: Multiple Risk Behaviour Index.

**Figure 4:**
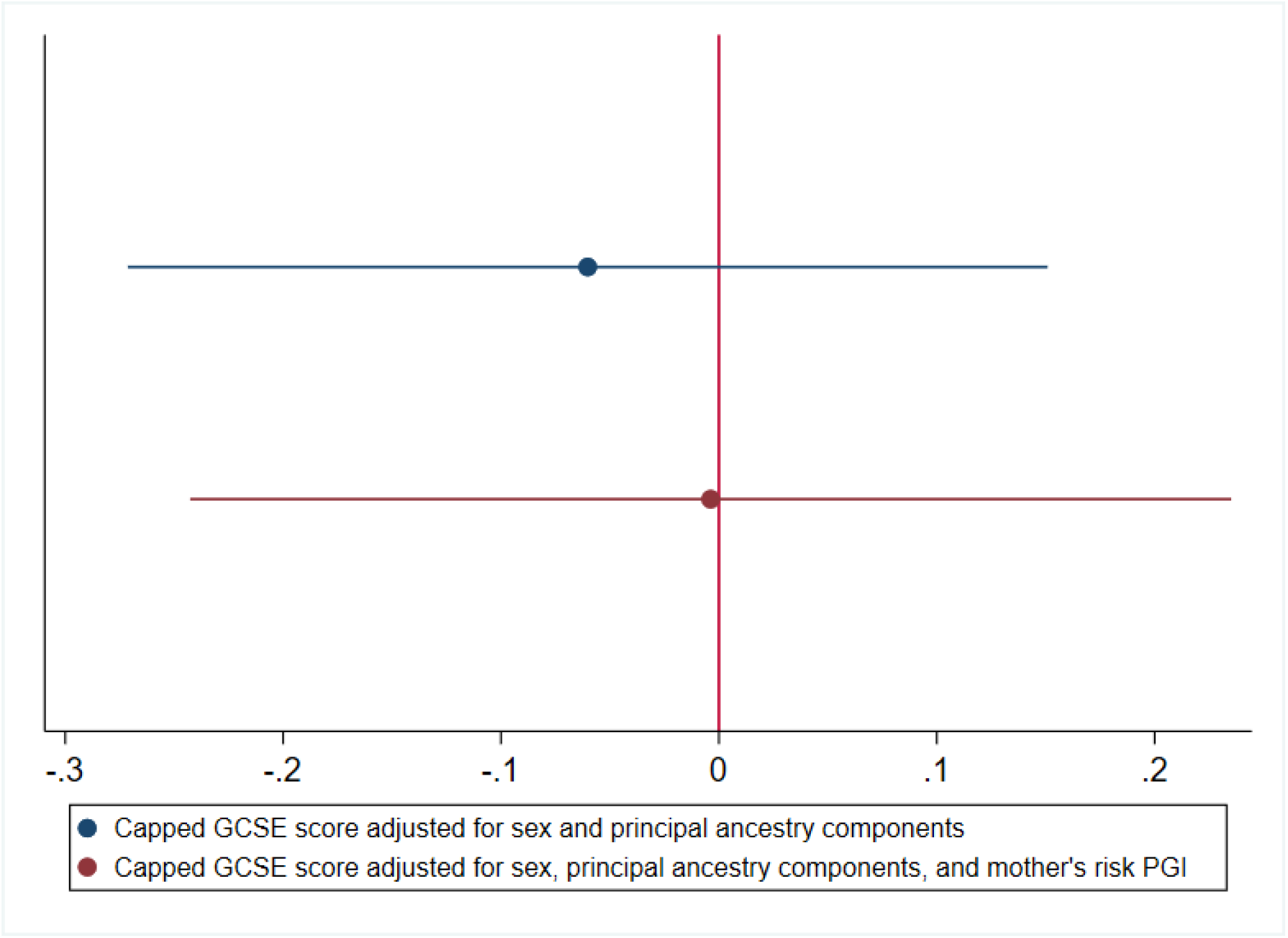
Association between the young person’s genetically-instrumented risk behaviour and their educational achievement (GCSE capped points score, standardized).

Figure 5 shows the association between the young person’s genetically instrumented capped GCSE score and the MRB index. There was a negative association between genetically instrumented education and the risk behaviours index (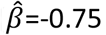, 95% CI: [-0.97, -0.54]) when adjusting for the young person’s sex and principal components of ancestry, and when additionally adjusting for the maternal educational achievement PGI (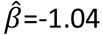, 95% CI: [-1.41, -0.67]). Which decreased the results for the binary outcome were similar (Supplementary Figure 3).

**Figure 5:**
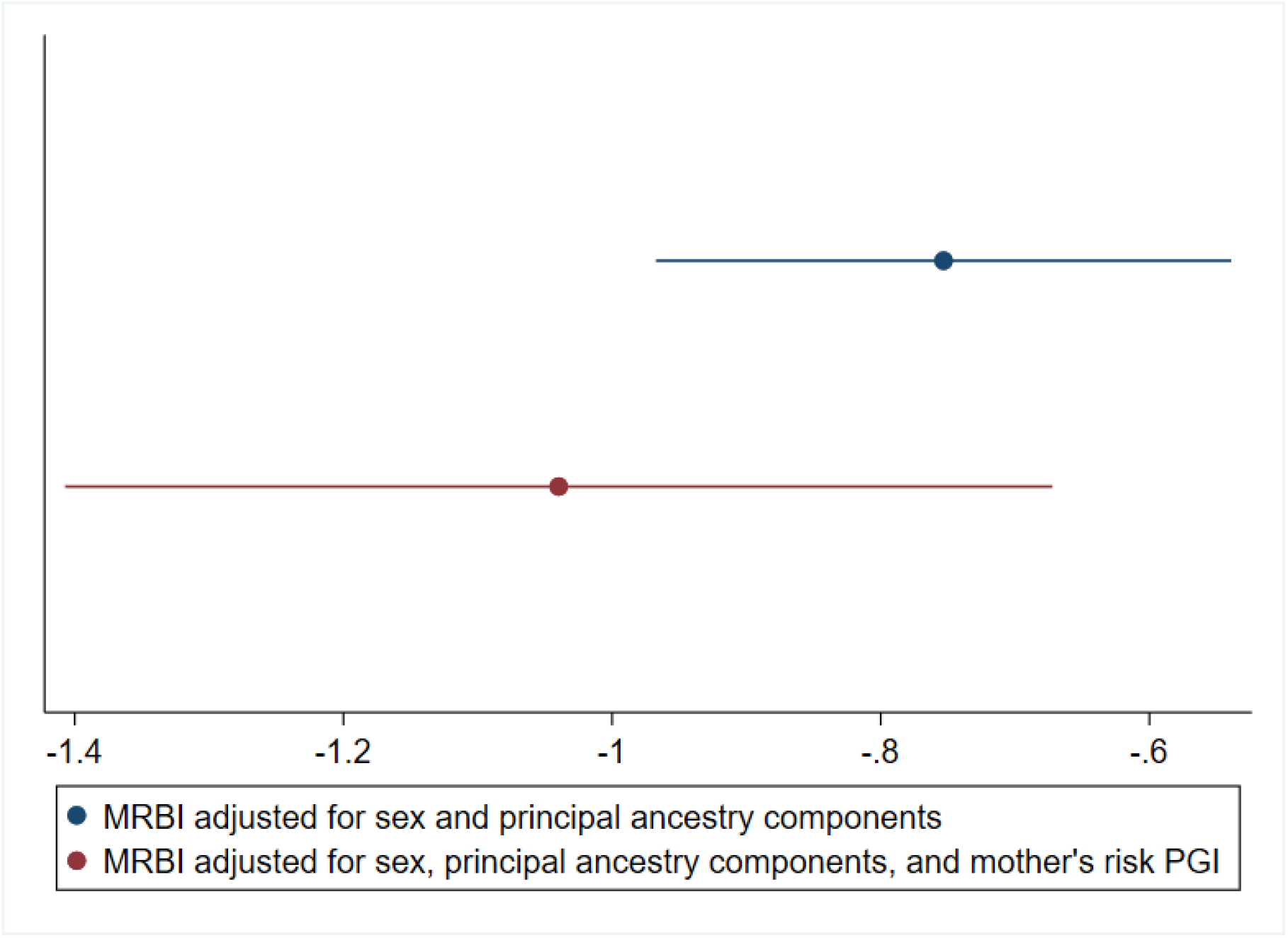
Association between the young person’s genetically-instrumented educational attainment (capped GCSE points score, standardized), and their Multiple Risk Behaviours (MRB) Index.

## Discussion

In a cohort of adolescents, an index of multiple risk behaviours was phenotypically associated with educational achievement at 16 after adjustment for confounders. Genetic analysis using GREML indicated not only that both traits are heritable, but also share genetic architecture, with considerable genetic overlap between the two traits. Consistent with results of phenotypic models, genetic variation associated with risk behaviours was negatively associated with educational achievement. Furthermore, results of bidirectional MR indicate that the principal direction of causation is likely to be from liability to educational achievement to risk behaviours. This suggests that engagement in risk behaviours may be partly driven by the education of an individual, or a closely related trait. In contrast, there was little evidence that genetic variation associated with engagement in risk behaviours causally affected educational achievement.

An alternative explanation for our results is familial factors, such as dynastic effects of parents on their children. Genetic nurture can occur when the parents’ genetic variants affect the offspring through environmental (i.e., non-transmitted) mechanisms. For example, Kong et al. (30) found that parents’ non-transmitted polygenic scores had an estimated association with the educational achievement of their children that was 29.9% (P= 1.6×10-14) of that of the transmitted polygenic score. (30) This is consistent with results found in Howe et al. (2022) within-sibship GWAS were educational attainment and phenotypes from population estimates, such as BMI and smoking, may be inflated by indirect genetic effects. However, adjusting for mothers’ polygenic score only modestly attenuated the effects. This attenuation could indicate that the effects are due to dynastic effects. However, to inferred dynastic effect in association genotyped mother father-children’s trios information is necessary. (16)

In our phenotypic analysis, we showed a negative association between MRB index and educational achievement for both educational achievement measures. We showed a decrease in capped GCSE score of -0.14 when the individual engaged in an additional risk behaviour; these results were slightly attenuated in the full model when controlled for confounders. The fully adjusted model showed a negative association in capped GCSES score of -0.11, which would represent a reduction of one grade in GCSE examination, for each additional risk behaviour the adolescent engages in. Similar results were observed when exploring the association between the MRB index and the probability of gaining A*-C GCSE including English and Mathematics. These results are consistent with previous results found in the ALSPAC cohort, where there was a negative association of multiple risk behaviours with education achievement of -6.31 (95% CI: [-7.03, -5.58]). (6) However, phenotypic associations are prone to reverse causation and confounding by characteristics not explained by covariates. Thus, we extended our analyses to a genetic causal inference approach to further explore this relationship.

The current literature gives similar estimates of the genetic heritability of educational achievement to those that we present. Among many others, Rimfeld et al. (32) found heritability for educational outcomes of 0.21 for GCSE Mathematics, 0.15 for GCSE English and 0.17 for GCSE Science. Likewise, Petrill et al. (33) found a heritability of reading performance of 0.38 in a genetic study using data from the Western Reserve Reading Project in Ohio, USA. Krapohl and Plomin (34) found a heritability of educational attainment of 0.31 in their study of socioeconomic position and offspring educational achievement. Our second set of results indicate that engagement in risk behaviours is strongly negatively associated with educational outcomes at 16 years. This effect was visible throughout the models that we tested, even when controlling for relevant confounders. The fully adjusted models showed a genetic correlation of -0.49 (pval=0.003) for our Capped GCSE scores and -0.75 (pval=0.005) for attaining 5 or more A*-C grades in Mathematics and English.

The Mendelian randomization analyses showed that genetic liability of adolescents for the MRB Index was not associated with educational achievement. Even when additionally adjusted for maternal risk PGI, the association was still null. This might suggest, along with our phenotypic results that there is a strong environmental factor driving the association between engagement in risk behaviours and educational achievement. Furthermore, our bidirectional MR suggested a causal effect of liability to educational attainment to engagement in risk behaviours. However, the reverse Mendelian Randomization estimates from risk behaviours to educational attainment had far lower power. This is because the risk behaviours PGI is weaker than the education PGI.

The risk behaviour literature shows that our considered behaviours co-occur frequently and also tend to cluster during adolescence (18,35). However, the current studies of clustered risk behaviour association focus on small-subset behaviours, such as alcohol use and smoking (36) and fail to account for a wider range of behaviours such as self-harm and criminal behaviour. Importantly, our analysis uses a wider range of clustered risk behaviours that allows us to capture a more complete effect on educational outcomes than seen in the existing literature. These insights support current policy studies’ recommendations that found intervention on clustering behaviour is more successful. (37) Furthermore, the associations found between multiple risk behaviour and education provides support to the current literature that shows school-based intervention have a higher rate of success (38); this could improve student outcomes and lessen the burden on public health services.

However, there are some limitations to our analysis. Missing data on risk behaviours and confounders may have reduced power and introduced bias, especially for GCTA analysis which did not use imputed data. Likewise, although the multiple risk behaviour score comprised a wide range of behaviours, by assigning each risk behaviour the same weight, we assumed that all risk behaviours contribute equally to associations with educational achievement. Our results also indicated the associations might be biased due to horizontal pleiotropy, where engagement in risk behaviour might be driven by a person’s education or other environmental factors. This is also observed in recent literature, where educational attainment drives certain risk behaviours, such as alcohol dependence (39) and smoking. (40) Lastly, our results might be confounded due to dynastic effects from the fathers. Our results controlling for the genotypes of the mothers were attenuated, but we did not have data on the genotypes of the fathers. Furthermore, some of the risk behaviours were measured via questionnaires, which makes the analysis subject to recall bias and desirability bias, where participants might have underreported socially perceived undesirable behaviours. Future work could investigate whether some risk behaviours are more closely linked to education than others. Our study only investigated the association of these phenotypes with common genetic variation, so future studies could investigate the impact of rare genetic variation.

In summary, we explored the genetic architecture of risk behaviour engagement in educational achievement and the bidirectional effect of these traits. We found that while liability to educational achievement was associated with fewer risk behaviours, there was little evidence that risk behaviours affected educational attainment. This suggests that educational achievement may be an effective intervention target for risky behaviours but not vice versa.

## Data Availability

Data from the ALSPAC cohort is available upon request and approval of the ALSPAC team. Applications can be made following these guidelines: http://www.bristol.ac.uk/alspac/researchers/access/

## Acknowledgement

We are extremely grateful to all the families who took part in this study, the midwives for their help in recruiting them, and the whole ALSPAC team, which includes interviewers, computer and laboratory technicians, clerical workers, research scientists, volunteers, managers, receptionists, and nurses. The UK Medical Research Council and Wellcome (Grant ref: 217065/Z/19/Z) and the University of Bristol provide core support for ALSPAC. This publication is the work of the authors and will serve as guarantors for the contents of this paper. A comprehensive list of grants funding is available on the on the ALSPAC website(http://www.bristol.ac.uk/alspac/external/documents/grant-acknowledgements.pdf). This research is funded by The Medical Research Council (MRC) and the University of Bristol support the MRC Integrative Epidemiology Unit [MC_UU_12013/1, MC_UU_12013/9, MC_UU_00011/1]. GWAS data was generated by Sample Logistics and Genotyping Facilities at Wellcome Sanger Institute and LabCorp (Laboratory Corporation of America) using support from 23andMe. The funders had no role in study design, data collection and analysis, decision to publish, or preparation of the manuscript.

## Code availability

All the code used to clean and analyse the data for this study is available: https://github.com/MichelleSpano/Risk-behaviours

## References

1. Viner RM, Ozer EM, Denny S, Marmot M, Resnick M, Fatusi A, et al. Adolescence and the social determinants of health. The Lancet. 2012;379(9826):1641–52.

2. Teh CH, Teh MW, Lim KH, Kee CC, Sumarni MG, Heng PP, et al. Clustering of lifestyle risk behaviours and its determinants among school-going adolescents in a middle-income country: a cross-sectional study. BMC Public Health [Internet]. 2019 Aug 27 [cited 2022 Oct 5];19(1). Available from: /pmc/articles/PMC6712662/

3. Mukamal KJ, Chiuve SE, Rimm EB. Alcohol Consumption and Risk for Coronary Heart Disease in Men With Healthy Lifestyles. Arch Intern Med. 2006 Oct 23;166(19):2145.

4. Li H, Khor CC, Fan J, Lv J, Yu C, Guo Y, et al. Genetic risk, adherence to a healthy lifestyle, and type 2 diabetes risk among 550,000 Chinese adults: results from 2 independent Asian cohorts. Am J Clin Nutr. 2020 Mar 1;111(3):698–707.

5. Gardner LA, Champion KE, Parmenter B, Grummitt L, Chapman C, Sunderland M, et al. Clustering of Six Key Risk Behaviors for Chronic Disease among Adolescent Females. Int J Environ Res Public Health. 2020 Oct 2;17(19):7211.

6. Wright C, Kipping R, Hickman M, Campbell R, Heron J. Effect of multiple risk behaviours in adolescence on educational attainment at age 16 years: a UK birth cohort study. BMJ Open. 2018 Jul 30;8(7):e020182.

7. Schuit AJ, van Loon AJM, Tijhuis M, Ocké MC. Clustering of lifestyle risk factors in a general adult population. Prev Med (Baltim). 2002;35(3):219–24.

8. Meader N, King K, Moe-Byrne T, Wright K, Graham H, Petticrew M, et al. A systematic review on the clustering and co-occurrence of multiple risk behaviours. BMC Public Health [Internet]. 2016 Jul 29 [cited 2022 Oct 5];16(1):1–9. Available from: https://bmcpublichealth.biomedcentral.com/articles/10.1186/s12889-016-3373-6

9. Brown JL, Gause NK, Northern N. The Association between Alcohol and Sexual Risk Behaviors among College Students: A Review. Curr Addict Rep [Internet]. 2016 Dec 1 [cited 2022 Oct 5];3(4):349. Available from: /pmc/articles/PMC5123847/

10. Bellis MA, Hughes K, Calafat A, Juan M, Ramon A, Rodriguez JA, et al. Sexual uses of alcohol and drugs and the associated health risks: A cross sectional study of young people in nine European cities. BMC Public Health. 2008 Dec 9;8(1):155.

11. Alamian A, Paradis G. Individual and social determinants of multiple chronic disease behavioral risk factors among youth. BMC Public Health. 2012 Dec 22;12(1):224.

12. Havdahl A, Hughes AM, Sanderson E, Ask H, Cheesman R, Reichborn-Kjennerud T, et al. Intergenerational effects of parental educational attainment on parenting and childhood educational outcomes: Evidence from MoBa using within-family Mendelian randomization. Available from: https://doi.org/10.1101/2023.02.22.23285699

13. Kipping RR, Smith M, Heron J, Hickman M, Campbell R. Multiple risk behaviour in adolescence and socio-economic status: findings from a UK birth cohort. Eur J Public Health. 2015 Feb;25(1):44–9.

14. Huesmann LR, Dubow EF, Boxer P. Continuity of aggression from childhood to early adulthood as a predictor of life outcomes: implications for the adolescent-limited and life-course-persistent models. Aggress Behav. 2009 Mar;35(2):136–49.

15. Mirza KAH, Mirza S. Adolescent substance misuse. Psychiatry. 2008 Aug;7(8):357–62.

16. Shore J, Janssen I. Adolescents’ engagement in multiple risk behaviours is associated with concussion. Inj Epidemiol. 2020 Dec 17;7(1):6.

17. MacArthur G, Caldwell DM, Redmore J, Watkins SH, Kipping R, White J, et al. Individual-, family-, and school-level interventions targeting multiple risk behaviours in young people. Cochrane Database of Systematic Reviews. 2018 Oct 5;2018(10).

18. Akasaki M, Ploubidis GB, Dodgeon B, Bonell CP. The clustering of risk behaviours in adolescence and health consequences in middle age. J Adolesc. 2019 Dec 1;77:188–97.

19. Boyd A, Tilling K, Cornish R, Davies A, Humphries K, MacLeod J. Professionally designed information materials and telephone reminders improved consent response rates: Evidence from an RCT nested within a cohort study. J Clin Epidemiol. 2015 Aug 1;68(8):877–87.

20. Fraser A, Macdonald-wallis C, Tilling K, Boyd A, Golding J, Davey smith G, et al. Cohort Profile: the Avon Longitudinal Study of Parents and Children: ALSPAC mothers cohort. Int J Epidemiol [Internet]. 2013 Feb [cited 2023 Mar 24];42(1):97–110. Available from: https://pubmed.ncbi.nlm.nih.gov/22507742/

21. Northstone K, Lewcock M, Groom A, Boyd A, Macleod J, Timpson N, et al. Open Peer Review The Avon Longitudinal Study of Parents and Children (ALSPAC): an update on the enrolled sample of index children in 2019 [version 1; peer review: 2 approved]. 2019 [cited 2023 Apr 6]; Available from: https://doi.org/10.12688/wellcomeopenres.15132.1

22. Teyhan A, Boyd A, Wijedasa D, MacLeod J. Early life adversity, contact with children’s social care services and educational outcomes at age 16 years: UK birth cohort study with linkage to national administrative records. BMJ Open. 2019 Oct 1;9(10).

23. Karlsson Linnér R, Biroli P, Kong E, Fleur Meddens SW, Wedow R, Alan Fontana M, et al. Genome-wide association analyses of risk tolerance and risky behaviors in over 1 million individuals identify hundreds of loci and shared genetic influences. [cited 2023 Mar 31]; Available from: https://doi.org/10.1038/s41588-018-0309-3

24. Smith GD, Ebrahim S. ‘Mendelian randomization’: can genetic epidemiology contribute to understanding environmental determinants of disease? Int J Epidemiol [Internet]. 2003 Feb [cited 2023 Mar 31];32(1):1–22. Available from: https://pubmed.ncbi.nlm.nih.gov/12689998/

25. Sanderson E, Davey Smith G, Bowden J, Munafò MR. Mendelian randomisation analysis of the effect of educational attainment and cognitive ability on smoking behaviour. Nat Commun. 2019 Dec 3;10(1):2949.

26. Brumpton B, Sanderson E, Heilbron K, Hartwig FP, Harrison S, Vie GÅ, et al. Avoiding dynastic, assortative mating, and population stratification biases in Mendelian randomization through within-family analyses. Nat Commun. 2020 Dec 1;11(1).

27. Davies NM, Holmes M V., Davey Smith G. Reading Mendelian randomisation studies: a guide, glossary, and checklist for clinicians. The BMJ [Internet]. 2018 [cited 2023 Mar 30];362. Available from: /pmc/articles/PMC6041728/

28. Okbay A, Wu Y, Wang N, Jayashankar H, Bennett M, Nehzati SM, et al. Polygenic prediction of educational attainment within and between families from genome-wide association analyses in 3 million individuals. Nat Genet. 2022 Apr 1;54(4):437–49.

29. Yang J, Lee SH, Goddard ME, Visscher PM. GCTA: A tool for genome-wide complex trait analysis. Am J Hum Genet. 2011 Jan 7;88(1):76–82.

30. Kong A, Thorleifsson G, Frigge ML, Vilhjalmsson BJ, Young AI, Thorgeirsson TE, et al. The nature of nurture: Effects of parental genotypes [Internet]. Available from: https://www.science.org

31. Morris TT, Davies NM, Hemani G, Smith GD. Population phenomena inflate genetic associations of complex social traits. Sci Adv [Internet]. 2020 Apr 1 [cited 2023 Mar 24];6(16). Available from: https://www.science.org/doi/10.1126/sciadv.aay0328

32. Rimfeld K, Kovas Y, Dale PS, Plomin R. Pleiotropy across academic subjects at the end of compulsory education. Scientific Reports 2015 5:1 [Internet]. 2015 Jul 23 [cited 2022 Oct 20];5(1):1–12. Available from: https://www.nature.com/articles/srep11713

33. Petrill SA, Hart SA, Harlaar N, Logan J, Justice LM, Schatschneider C, et al. Genetic and environmental influences on the growth of early reading skills. J Child Psychol Psychiatry. 2010 Jun;51(6):660–7.

34. Krapohl E, Plomin R. Genetic link between family socioeconomic status and children’s educational achievement estimated from genome-wide SNPs. Mol Psychiatry. 2016 Mar 1;21(3):437–43.

35. Hair EC, Park MJ, Ling TJ, Moore KA. Risky Behaviors in Late Adolescence: Co-occurrence, Predictors, and Consequences. Journal of Adolescent Health. 2009 Sep;45(3):253–61.

36. Bannink R, Broeren S, Heydelberg J, van’t Klooster E, Raat H. Depressive symptoms and clustering of risk behaviours among adolescents and young adults attending vocational education: a cross-sectional study. BMC Public Health. 2015 Dec 18;15(1):396.

37. Rabel M, Laxy M, Thorand B, Peters A, Schwettmann L, Mess F. Clustering of health-related behavior patterns and demographics. Results from the population-based KORA S4/F4 cohort study. Front Public Health. 2019 Jan 22;6(JAN):387.

38. Macarthur G, Caldwell DM, Redmore J, Watkins SH, Kipping R, White J, et al. Individual-, family-, and school-level interventions targeting multiple risk behaviours in young people. Cochrane Database Syst Rev [Internet]. 2018 Oct 5 [cited 2022 Oct 20];10(10). Available from: https://pubmed.ncbi.nlm.nih.gov/30288738/

39. Rosoff DB, Clarke TK, Adams MJ, Mcintosh AM, George •, Smith D, et al. Educational attainment impacts drinking behaviors and risk for alcohol dependence: results from a two-sample Mendelian randomization study with ∼780,000 participants. Mol Psychiatry [Internet]. 2021 [cited 2023 Mar 24];26:1119–32. Available from: https://doi.org/10.1038/s41380-019-0535-9

40. Latvala A, Rose RJ, Pulkkinen L, Dick DM, Korhonen T, Kaprio J. Drinking, smoking, and educational achievement: Cross-lagged associations from adolescence to adulthood. Drug Alcohol Depend [Internet]. 2014 [cited 2023 Mar 24];137(1):106–13. Available from: https://click.endnote.com/viewer?doi=10.1016%2Fj.drugalcdep.2014.01.016&token=WzM1MDQ0MzgsIjEwLjEwMTYvai5kcnVnYWxjZGVwLjIwMTQuMDEuMDE2Il0.T6uvzxLZclEkAgPJkUWjypmZE

