## Supplementary document for "Genetic associations of risk behaviours and educational attainment"

### Contents:

**Figure 1: STROBE diagram describing selection of sample and imputed sample.**

**Table 1: Descriptive statistics for MRBs in complete case sample.**

**Table 2: Comparison of observed characteristics between participants in the complete dataset (N=1,735) and incomplete dataset (N=15,616).**

**Table 3: Association of Capped GCSE score and score with an index of multiple risk behaviours, based on the complete case sample.**

**Table 4: Association of achieving five or more A*-C GCSE score with an index of multiple risk behaviours, based on the complete case sample.**

**Table 5: descriptive statistics for educational outcomes and exposure, MRB Index in complete case sample.**

**Table 6: Associations of probability of gaining 5 or more A*-C GCSEs including English and Maths with an index of multiple risk behaviours, based on imputed data (N=7,695).**

**Figure 2: Association between the young person's genetically instrumented multiple risk behaviours (MRB) index and the probability of gaining 5 or more A* - C GCSEs including English and Maths.**

**Figure 3: Association between the young person's genetically-instrumented educational attainment (obtaining attaining five or more A*-C GCSEs including English and Maths), and their Multiple Risk Behaviours (MRB) Index.**

Figure 1: STROBE diagram describing selection of sample and imputed sample.

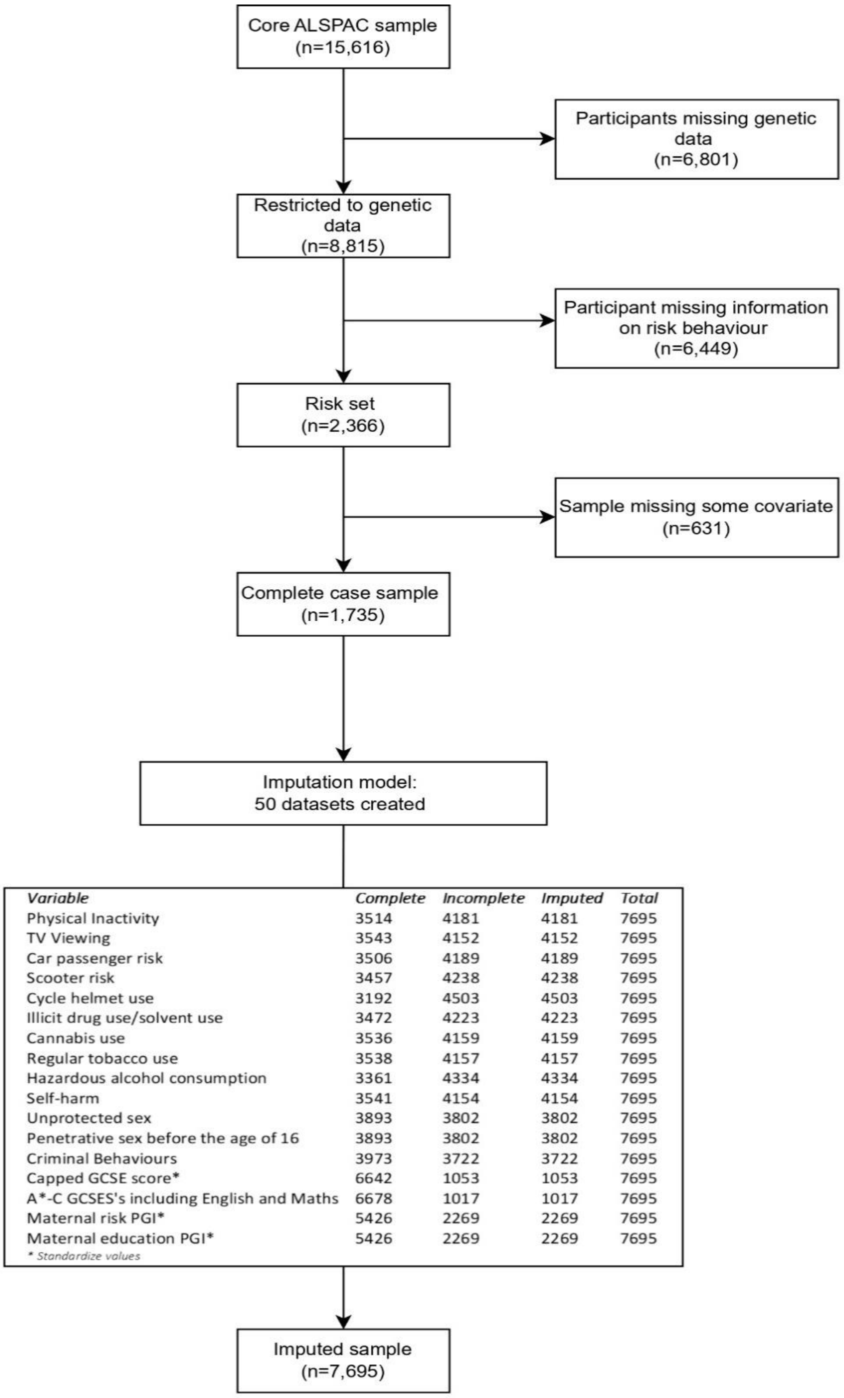

| *Variable* | *Complete* | *Incomplete* | *Imputed* | *Total* |
|---|---|---|---|---|
| Physical Inactivity | 3514 | 4181 | 4181 | 7695 |
| TV Viewing | 3543 | 4152 | 4152 | 7695 |
| Car passenger risk | 3506 | 4189 | 4189 | 7695 |
| Scooter risk | 3457 | 4238 | 4238 | 7695 |
| Cycle helmet use | 3192 | 4503 | 4503 | 7695 |
| Illicit drug use/solvent use | 3472 | 4223 | 4223 | 7695 |
| Cannabis use | 3536 | 4159 | 4159 | 7695 |
| Regular tobacco use | 3538 | 4157 | 4157 | 7695 |
| Hazardous alcohol consumption | 3361 | 4334 | 4334 | 7695 |
| Self-harm | 3541 | 4154 | 4154 | 7695 |
| Unprotected sex | 3893 | 3802 | 3802 | 7695 |
| Penetrative sex before the age of 16 | 3893 | 3802 | 3802 | 7695 |
| Criminal Behaviours | 3973 | 3722 | 3722 | 7695 |
| Capped GCSE score* | 6642 | 1053 | 1053 | 7695 |
| A*-C GCSES's including English and Maths | 6678 | 1017 | 1017 | 7695 |
| Maternal risk PGI* | 5426 | 2269 | 2269 | 7695 |
| Maternal education PGI* | 5426 | 2269 | 2269 | 7695 |

** Standardize values*

Table 1: Descriptive statistics for MRBs in complete case sample.

| Multiple Risk Behaviours (MRBs) | n | % Engaging |
|---|---|---|
| Physical Inactivity | 3556 | 74% |
| TV Viewing | 3584 | 21% |
| Car passenger risk | 3547 | 30% |
| Scooter risk | 3497 | 20% |
| Cycle helmet use | 3227 | 24% |
| Illicit drug use/solvent use | 3512 | 8% |
| Cannabis use | 3578 | 10% |
| Regular tobacco use | 3579 | 12% |
| Hazardous alcohol consumption | 3399 | 36% |
| Self-harm | 3582 | 19% |
| Penetrative sex before the age of 16 | 3933 | 17% |
| Unprotected sex | 3933 | 3% |
| Criminal Behaviours | 4017 | 47% |

Table 2: Comparison of observed characteristics between participants in the complete dataset (N=1,735) and incomplete dataset (N=15,616)

| | Complete dataset N=1,735 | Incomplete dataset N=15,616 | Test for difference |
|---|---|---|---|
| ***Continuous variables*** | ***Mean*** | ***Mean*** | ***T-test*** |
| MRB Index | 3.18 | 3.17 | -0.15 |
| Capped GCSE score | 375.28 | 304.06 | -31.58 |
| Maternal age | 29.72 | 27.71 | -16.95 |
| Cognitive ability | 109.09 | 102.05 | -16.51 |
| ***Categorical variables*** | ***%*** | ***%*** | ***Chi²*** |
| Achieved 5 or more A*- C GCSEs including English and maths | | | 739.55 |
| No | 21.66 | 54.86 | |
| Yes | 78.34 | 45.14 | |
| Sex | | | 76.73 |
| Male | 42.02 | 52.54 | |
| Female | 57.98 | 47.46 | |
| Maternal Education | | | 407.52 |
| <O level | 13.86 | 33.18 | |
| O level | 34.22 | 34.72 | |
| A level | 30.92 | 20.75 | |
| Degree | 21.01 | 11.35 | |
| Housing Tenure | | | 343.38 |
| Mortgage/owned | 89.94 | 70.28 | |
| Council rented | 3.90 | 16.20 | |
| Private/other rented | 6.15 | 13.52 | |
| Parent social class | | | 199.34 |
| Professional | 19.06 | 12.10 | |
| Managerial and technical | 48.27 | 40.41 | |
| Skilled non-manual | 22.46 | 26.12 | |
| Skilled manual | 10.21 | 21.37 | |

Table 3: Association of Capped GCSE score and score with an index of multiple risk behaviours, based on the complete case sample.

| **GCSE capped point score** | **Model 1**<br>**N = 2053** | **Model 2**<br>**N=1879** | **Model 3**<br>**N=1854** | **Model 4**<br>**N=1736** |
|---|---|---|---|---|
| | 95% confidence intervals in brackets; p-values in parentheses. | | | |
| **MRB Index** | -0.07 [-0.09, -0.05] | -0.06 [-0.07, -0.04] | -0.06 [-0.07, -0.04] | -0.06 [-0.07, -0.04] |
| **Parental socioeconomic Position** | | ✓ | ✓ | ✓ |
| **Maternal Education ref: <O level** | | ✓ | ✓ | ✓ |
| **Sex ref: male** | | ✓ | ✓ | ✓ |
| **Housing tenure ref: Owned** | | | ✓ | ✓ |
| **Cognitive ability** | | | | ✓ |

Table 4: Association of achieving five or more A*-C GCSE score with an index of multiple risk behaviours, based on the complete case sample.

| **A*-C GCSE's including English and Maths** | **Model 1**<br>**N=2053** | **Model 2**<br>**N=1879** | **Model 3**<br>**N=1854** | **Model 4**<br>**N=1736** |
|---|---|---|---|---|
| | Exponentiated Coefficients. 95% confidence intervals in brackets; p-values in parentheses. | | | |
| **MRB Index** | 0.86 [0.82, 0.91] | 0.87 [0.82, 0.91] | 0.83 [0.82, 0.92] | 0.86 [0.81, 0.91] |
| **Parental socioeconomic Position** | | ✓ | ✓ | ✓ |
| **Maternal Education ref: <O level** | | ✓ | ✓ | ✓ |
| **Sex ref: male** | | ✓ | ✓ | ✓ |
| **Housing tenure ref: Owned** | | | ✓ | ✓ |
| **Cognitive ability** | | | | ✓ |

Table 5: descriptive statistics for educational outcomes and exposure, MRB Index in complete case sample.

| Variable | N | Mean | SD | Min | Max |
|---|---|---|---|---|---|
| Capped GCSE score | 6654 | 329.67 | 89.56 | 0 | 540 |
| A*-C GCSES's including English and Maths | 6695 | 0.57 | 0.50 | 0 | 1 |
| MRB Index | 2171 | 3.19 | 1.97 | 0 | 11 |

Table 6: Associations of probability of gaining 5 or more A*-C GCSEs including English and Maths with an index of multiple risk behaviours, based on imputed data (N=7,695)

| **A*-C GCSEs including English and Maths** | **Model 1** | **Model 2** | **Model 3** | **Model 4** |
|---|---|---|---|---|
| | Exponentiated Coefficients. 95% confidence intervals in brackets; p-values in parentheses. | | | |
| **MRBS** | 0.81 [0.78, 0.84] | 0.82[0.79, 0.86] | 0.83 [0.80, 0.86] | 0.81 [0.77, 0.84] |
| **Parental socioeconomic Position** | | ✓ | ✓ | ✓ |
| **Maternal Education ref: <O level** | | ✓ | ✓ | ✓ |
| **Sex ref: male** | | ✓ | ✓ | ✓ |
| **Housing tenure ref: Owned** | | | ✓ | ✓ |
| **Cognitive ability** | | | | ✓ |

Figure 2: Association between the young person's genetically instrumented multiple risk behaviours (MRB) index and the probability of gaining 5 or more A* - C GCSEs including English and Maths.

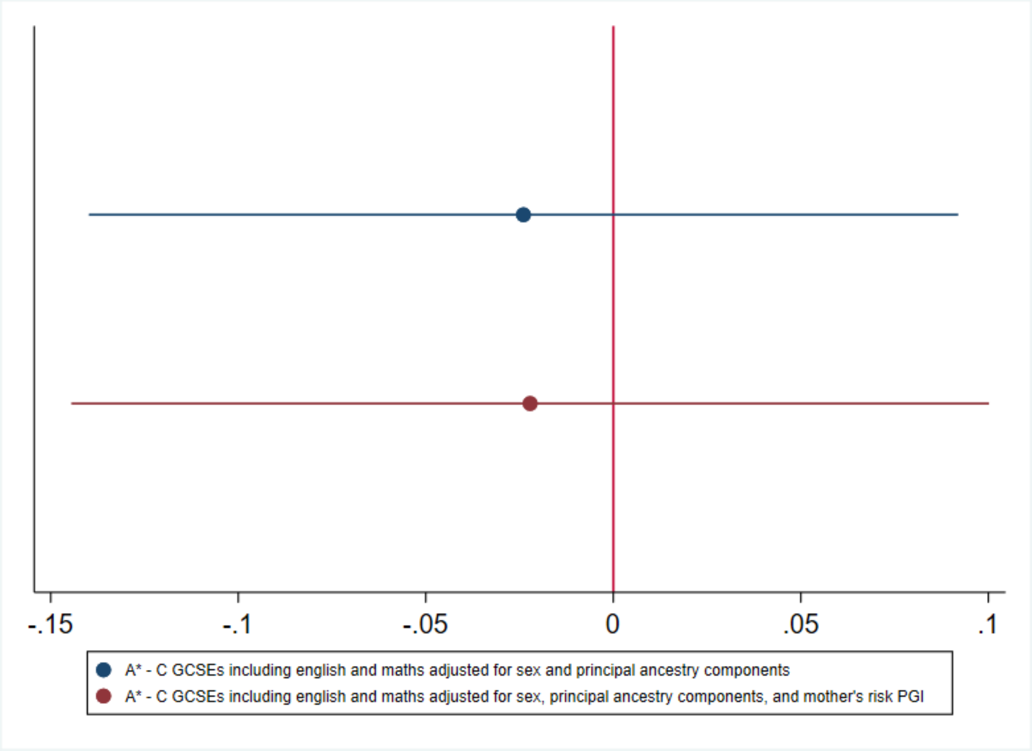

Figure 3: Association between the young person's genetically-instrumented educational attainment (obtaining attaining five or more A*-C GCSEs including English and Maths), and their Multiple Risk Behaviours (MRB) Index.

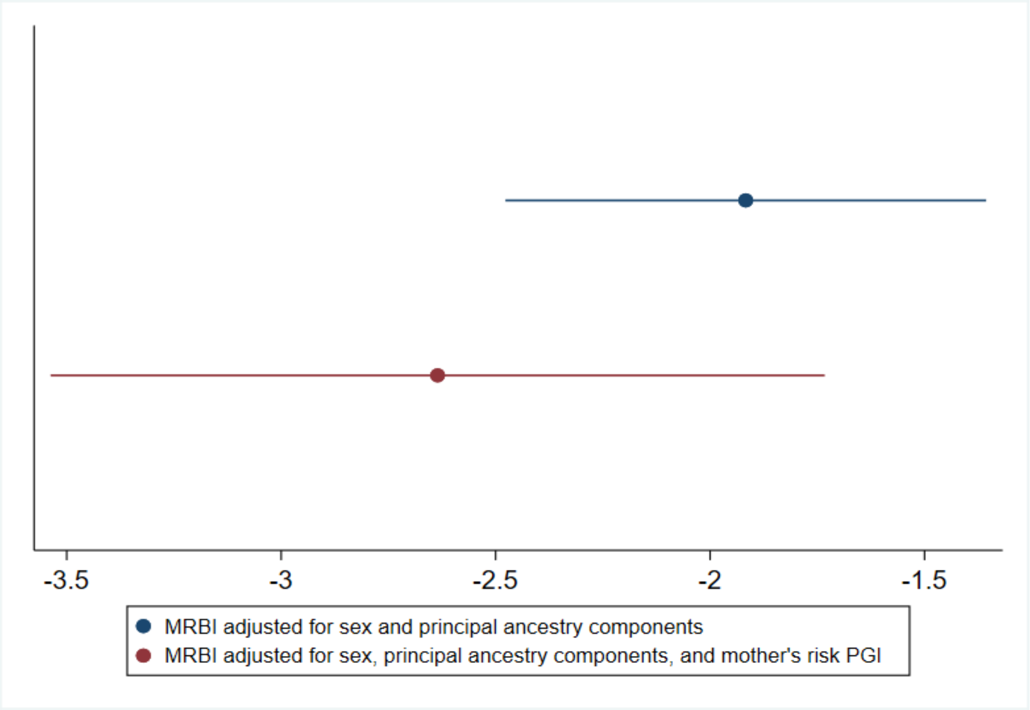
